# Sotatercept Improves Small Airway Disease and Hyperinflation in Patients with Pulmonary Hypertension

**DOI:** 10.1101/2025.10.08.25337393

**Authors:** Hooman D. Poor, Jimmy Zhang, Simone Hannah-Clark, Shyla Saini, Elliot Eisenberg, Jing Wang, Alison G. Lee, Gregory Serrao, Helena Schotland, Linda Rogers, Benjamin M. Smith, Mary Beth Beasley, Michelle Li, Charles A. Powell, Corey E. Ventetuolo, Maria Padilla

## Abstract

**Rationale:** Small airways disease (SAD) and hyperinflation are common in precapillary pulmonary hypertension (PH). Activin signaling plays an important role in airway and bronchial function.

**Objective:** To determine whether treatment with sotatercept, an activin signaling inhibitor, for severe precapillary PH is associated with improvements in physiologic markers of SAD and hyperinflation.

**Methods:** We conducted a single-center, retrospective cohort study of participants who received sotatercept for the treatment of severe precapillary PH despite background PH treatments who also had pulmonary function tests (PFT) before and after initiation of sotatercept treatment.

**Measurements and Main Results:** Forty-eight participants were included (median age 68 years, 77% female). Median BMI was 26.7 kg/m^2^ (IQR 23.6-31.4). All participants were functional class III or IV. Follow-up PFTs obtained a median of 4.4 months after sotatercept initiation showed significant improvements: FEV1 +155 mL (11%, 95% confidence interval [CI], 100-215 mL; p<0.001), FVC +180 mL (10%, 95% CI, 125-245 mL; p<0.001), FEF25-75% +0.15 L/sec (16%, 95% CI, 0.03-0.28 L/sec; p=0.015), DLCO +0.79 mL/min/mmHg (10%, 95% CI, 0.30-1.25 mL/min/mmHg; p<0.01). In participants with paired lung volume measurements (n=22), RV decreased 210 mL (12%, 95% CI, -340 to -85 mL; p<0.01), RV/TLC decreased 5% (95% CI, -7% to -3%; p<0.001), and ERV increased 175 mL (29%, 95% CI, 50-385 mL; p=0.02). There was no overall change in TLC or FRC.

**Conclusion:** In a real-world cohort of patients with severe precapillary PH from a variety of causes, sotatercept was associated with improvements in markers of SAD and hyperinflation.

## INTRODUCTION

Pulmonary arterial hypertension (PAH) is a form of precapillary pulmonary hypertension (PH) characterized by pulmonary vascular remodeling [1]. Patients with PAH are noted to have high rates of small airways disease (SAD) [2–4], and some patients with advanced PAH display progressive obstructive airways physiology [5]. Dynamic hyperinflation, a consequence of SAD and obstructive airways disease, contributes to exercise intolerance and dyspnea in patients with PAH [6]. Notably, “PAH with lung phenotype” has recently been described as a subtype among idiopathic PAH patients that occurs more frequently in older male patients who have a significant tobacco history, low DLCO, and marked hypoxemia in the absence of parenchymal lung disease [7, 8]. The mechanisms and implications of SAD and obstructive airways disease in PAH are not entirely clear.

Precapillary PH and combined pre- and post-capillary PH (CpcPH) frequently occurs in cardiopulmonary conditions other than PAH, such as left heart failure, interstitial lung disease (ILD), chronic obstructive pulmonary disease (COPD), and sarcoidosis [9]. These other cardiopulmonary conditions are also associated with significant rates of SAD, the presence of which is associated with worse exercise capacity and increased mortality [10–14]. In the real world, significant precapillary PH can occur with comorbid cardiopulmonary diseases and patients can have overlapping clinical classifications, as well as “phenotypic drift” from PAH to CpcPH [15, 16]. PAH with comorbidities has emerged as a specific consideration in recent guidelines and off-label use of PAH medications can occur in this setting, especially in clinically dire circumstances [9, 17]. Cassady and Gladwin recently asserted that vulnerable and understudied PH patients, such as those with precapillary PH due to sarcoidosis and sickle cell disease, should have access to PAH pharmacotherapy under the banner of individualized therapy [18].

Sotatercept, an activin-signaling inhibitor, is a highly effective therapy approved for the treatment of PAH and being studied in CpcPH [19–21]. The purported mechanism for sotatercept’s efficacy in PAH is the rebalancing of proliferative and antiproliferative pathways in the pulmonary arterioles [22]. We previously reported improved lung function, and in particular small airway function, with sotatercept in patients with sarcoidosis-associated PH (in press). The aim of this study was to assess changes in lung function, specifically markers of SAD and hyperinflation, in a broader group of patients with significant precapillary PH treated with sotatercept. We hypothesized that markers of SAD and hyperinflation would improve in patients with precapillary PH after treatment with sotatercept.

## METHODS

### Participants and Clinical Data

We retrospectively studied patients followed in a pulmonary vascular disease clinic at a single academic medical center who received sotatercept for the treatment of significant precapillary PH from April 2024 to August 2025. The Mount Sinai Institutional Review Board gave ethical approval for this work (IRB approval 19-00263). Any patient who received sotatercept for the treatment of precapillary PH outside of the indication of classical PAH was clinically counseled about the off-label use of sotatercept.

Patients were included in the study if they had pulmonary function testing (PFT) done both before and after the initiation of sotatercept, no more than two years apart. Participant data, including age, biological sex, height, weight, medical comorbidities, concomitant pulmonary vasodilator use, and functional class (FC) were extracted from the electronic medical record. While all participants had severe precapillary PH, right ventricular dysfunction, and/or significant exercise intolerance necessitating further treatment, for the purposes of this report participants were categorized into PH subgroups based on the diagnostic work up for their initial PH diagnosis as per the standard World Symposium on PH (WSPH) classification system [9].

### Pulmonary Function Testing

PFT data were collected from the electronic medical record. All participants included in the study had completed spirometry pre- and post-sotatercept treatment. Body plethysmography and diffusing capacity, if available, were also collected. If participants had multiple PFTs prior to initiation of sotatercept, the PFTs closest to the sotatercept start date were used. If participants had multiple PFTs after the initiation of sotatercept, the most recent PFT that was completed within two years of baseline (the pre-sotatercept PFT) was used.

### Hemodynamic Data

Right heart catheterization (RHC) data prior to sotatercept initiation was available for all participants (the majority occurring within one year of sotatercept initiation). RHC data after sotatercept initiation was available for some participants. Mean pulmonary artery pressure (mPAP), systolic PAP (sPAP), diastolic PAP (dPAP), right atrial pressure (RAP), pulmonary capillary wedge pressure (PCWP), and heart rate (HR) during the procedure were collected. For subjects with cardiac output (CO) measured by both indirect Fick and thermodilution methods, values acquired by thermodilution were used; indirect Fick values were used if thermodilution was not performed. Pulmonary vascular resistance (PVR) was calculated using the standard equation of (mPAP – PCWP)/CO, expressed in Wood units. Stroke volume (SV) was calculated as CO/HR. Pulmonary artery compliance (PAC) was calculated as SV/(sPAP-dPAP).

### Statistical Analysis

Pre- and post-sotatercept measurements were compared using Wilcoxon signed-rank tests with continuity correction. Pseudo-medians estimate the median of the difference between pre-sotatercept and post-sotatercept measurements. In addition, the exact p-value and confidence intervals are calculated when the differences contain no zeroes or ties; otherwise, a normal approximation is used. Associations between changes in lung volumes were evaluated with simple linear regression. A *p* value less than 0.05 was considered statistically significant. All analyses were performed using GraphPad Prism 10 and R version 4.3.1 (stats package).

## RESULTS

### Baseline Characteristics

Forty-eight participants were included in the study. Participants had a median age of 68 years (interquartile range [IQR], 58-77). Thirty-seven participants (77%) were female and the median body mass index was 26.7 kg/m^2^ (IQR 23.6-31.4). Seventeen (35%) participants had PAH, five (10%) had Group 2 PH (all with CpcPH secondary to heart failure with preserved ejection fraction [HFpEF]), 12 (25%) had Group 3 PH (11 with ILD and one with COPD), and 14 (29%) had sarcoidosis-associated PH. The five participants with HFpEF-PH had significant precapillary PH and severe right ventricular dysfunction on echocardiogram. All participants were FC III or IV. Demographics, comorbidities, background pulmonary vasodilator therapy, as well as baseline hemodynamics, lung function, and FC are listed in Table 1.

**Table 1:**
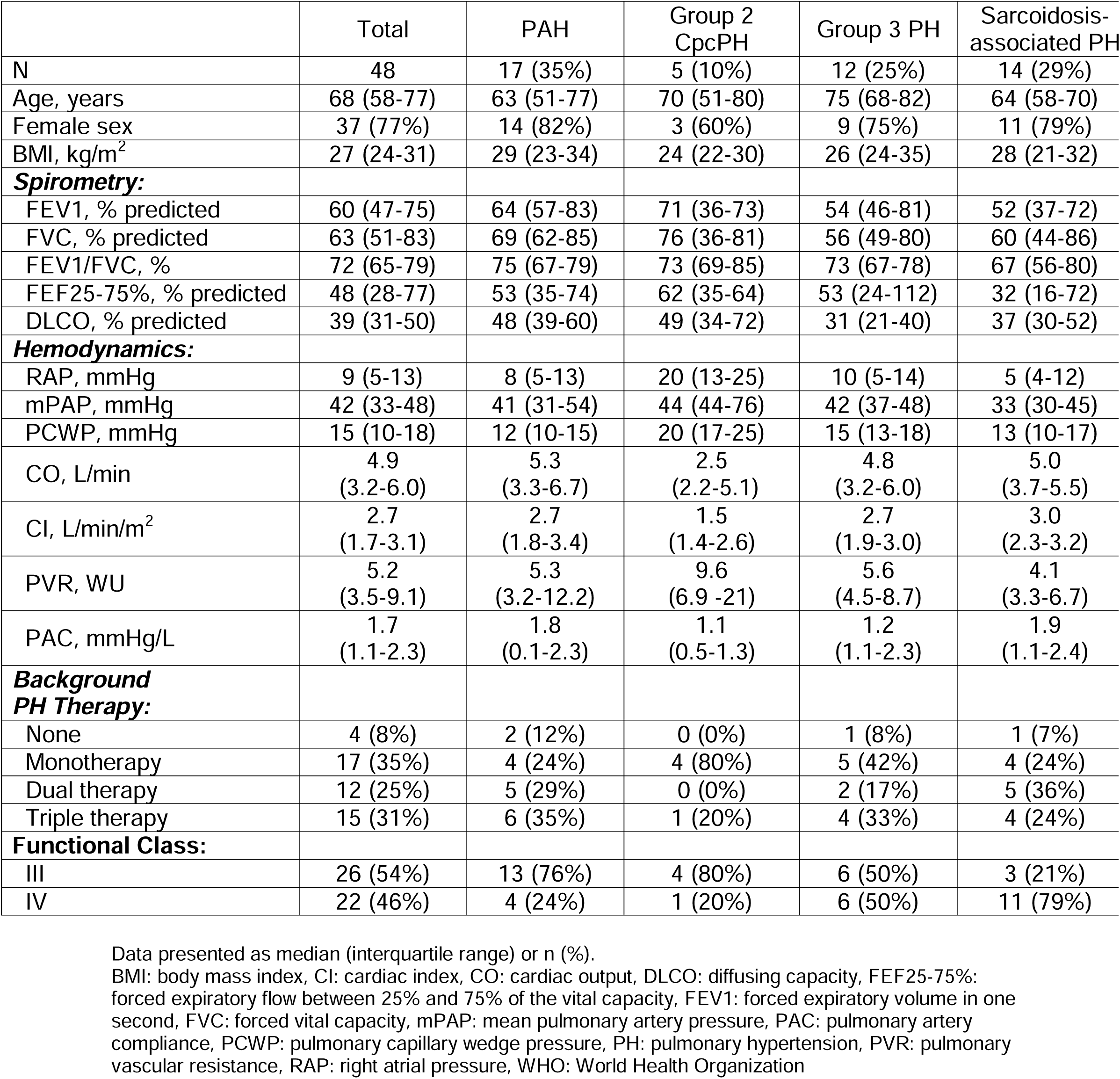
Baseline Characteristics.

Baseline body plethysmography was available in 27 participants and summarized in Table 2. Three participants (11%) had total lung capacity (TLC) greater than 100% predicted, 12 (44%) had residual volume (RV) greater than 100% predicted, and 20 (74%) had RV/TLC greater than 100% predicted.

**Table 2:** Baseline Lung Volumes.

|  | Total | PAH | Group 2<br>Cpc-PH | Group 3<br>PH | Sarcoidosis-associated PH |
| --- | --- | --- | --- | --- | --- |
| N | 27 (56%) | 12 (71%) | 3 (60%) | 6 (50%) | 6 (43%) |
| TLC, % predicted | 72<br>(63-89) | 79<br>(66-97) | 81<br>(50-89) | 68<br>(50-84) | 66<br>(47-82) |
| FRC, % predicted | 90<br>(66-103) | 93<br>(68-103) | 93<br>(73-96) | 76<br>(54-94) | 95<br>(58-109) |
| RV, % predicted | 91<br>(67-114) | 105<br>(83-125) | 89<br>(81-142) | 64<br>(52-87) | 91<br>(57-156) |
| RV/TLC, %<br>predicted | 131<br>(98-149) | 133<br>(110-140) | 162<br>(103-183) | 91<br>(82-121) | 141<br>(82-230) |
| ERV, % predicted | 69<br>(45-96) | 69<br>(45-89) | 46<br>(33-134) | 67<br>(55-121) | 68<br>(45-122) |

Data presented as median (interquartile range) or n (%)
ERV: expiratory reserve volume, FRC: functional residual capacity, RV: residual volume, SAPH: sarcoidosis-associated pulmonary hypertension, TLC: total lung capacity, WHO: World Health Organization

### Spirometry and Diffusing Capacity

The median time between pre- and post-sotatercept PFTs was 9.4 months (IQR 3.3-17.3). Multiple pulmonary function parameters significantly improved following the initiation of sotatercept (median time 4.4 months, IQR 2.0-7.8)(Figure 1). FEV1 increased in 40 (83%) participants (median increase 155 mL [11%], 95% confidence interval [CI], 100-215 mL; p<0.001) and FVC increased in 40 (83%) participants (median increase 180 mL [10%], 95% CI, 125-245 mL; p<0.001). Median FEF25-75% increased 0.15 L/sec (16%; 95%CI, 0.03-0.28 L/sec; p=0.015). There was no difference in peak expiratory flow (PEF) or FEV1/FVC. Notably, 23 participants (48%) had a decline in FEV1/FVC, of which 20 (87%) had significant increases in FVC (median FVC change 225 mL, 95% CI, 150-340; p<0.001), and FEV1 (median FEV1 increase 95 mL, 95% CI, 40-160; p=0.003). FEF25-75% decreased in 19 participants, of which 15 (79%) had increases in FVC (median FVC increase 170 mL, 95% CI, 80-285; p=0.001). In the 36 (75%) participants who had DLCO measurements both pre-and post sotatercept, DLCO increased 0.79 mL/mmHg/min (10%, 95% CI, 0.30-1.25 mL/mmHg/min; p<0.01).

**Figure 1:**
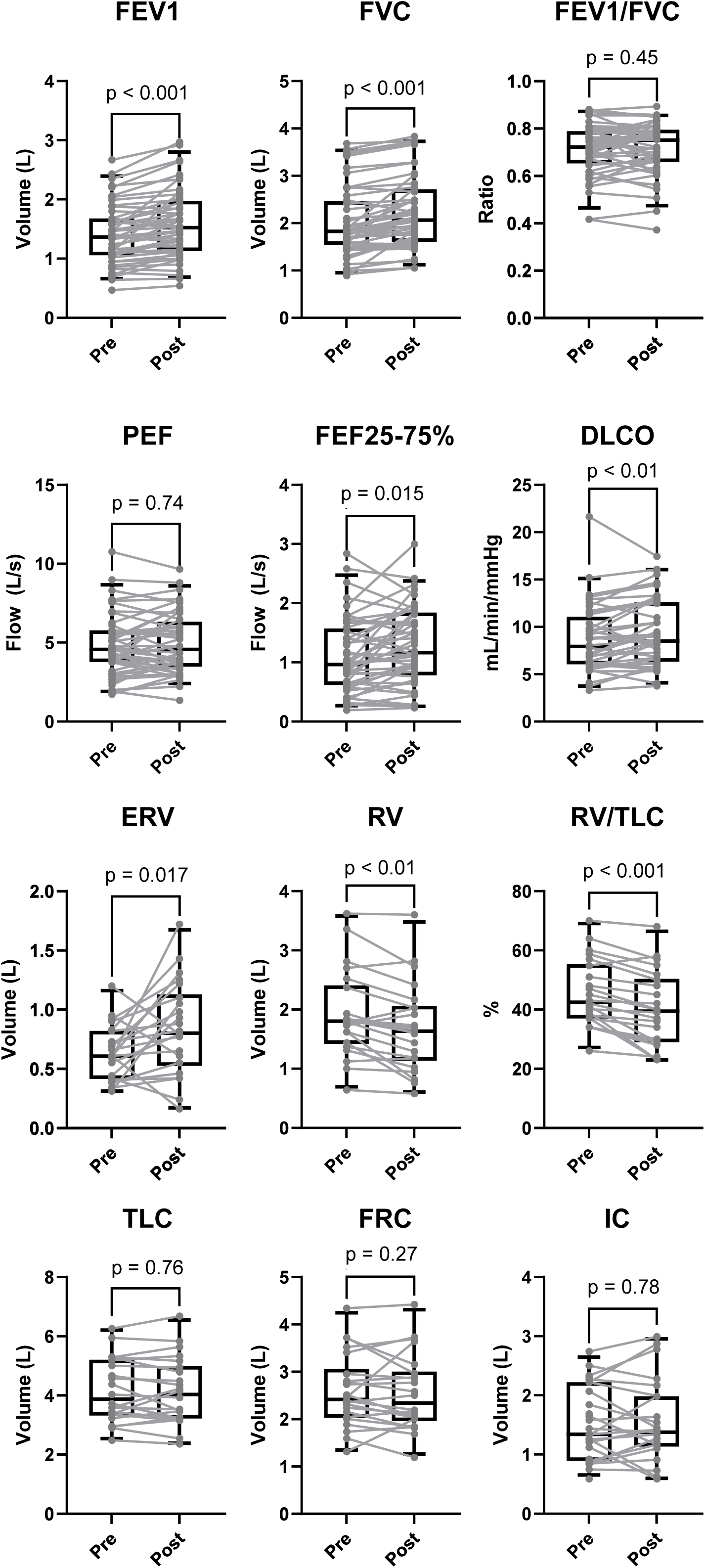
Changes in spirometry, lung volumes, and diffusing capacity before and after initiation of sotatercept. There were significant increases in FEV1, FVC, FEF25-75%, DLCO, and ERV. There were significant decreases in RV and RV/TLC. DLCO: diffusing capacity, ERV: expiratory reserve volume, FEF25-75%: forced expiratory flow between 25% and 75% of the vital capacity, FEV1: forced expiratory volume in one second, FVC: forced vital capacity, FRC: functional residual capacity, IC: inspiratory capacity, RV: residual volume, TLC: total lung capacity

### Lung Volumes

Twenty-two (46%) participants had body plethysmography both pre and post-sotatercept (Table 2). RV decreased in 18 (82%) participants (median decrease 210 mL [12%], 95% CI, -340 to -85 mL; p<0.01), and RV/TLC ratio decreased in 21 (95%) participants (median decrease 5%, 95% CI, -7% to -3%; p<0.001). Expiratory reserve volume (ERV) increased 175 mL (29%, 95% CI, 50-385 mL; p=0.02). While overall there was no difference in TLC between both groups, TLC decreased in 14 participants (64%). In participants whose TLC decreased, there was also a marked reduction in RV (median RV change -340 mL, 95% CI, -490 to -220; p=0.001). For the 8 (36%) participants with an increase in TLC, there was no significant change in RV (median RV change +20 mL, 95% CI, -130 to 185; p=0.74) [Figure S1A]. Eleven (50%) participants had increases in FRC, all of whom had increases in ERV (median ERV increase 390 mL, 95% CI, 100-670; p<0.01), whereas there was no difference in ERV when FRC decreased (median change in ERV 53 mL, 95% CI, -180 to 285; p=0.62) [Figure S1B]. Changes in TLC correlated with changes in RV and changes in in FRC correlated with changes in ERV. There was no relationship between changes in ERV and changes in RV, TLC, or FVC (Figure S2).

### Hemodynamics

Repeat hemodynamics were performed in 23 participants (Figure S3). There were significant improvements in RAP (median change -4 mmHg, 95% CI -7 to 0; p=0.04), mPAP (median change -14 mmHg, 95% CI, -18 to -10; p<0.001), PVR (median change -3.0, 95% CI, -7.0 to -1.4; p<0.001), and PAC (median change 0.60, 95% CI, 0.29-0.92; p<0.001). There was a trend towards a decrease in PCWP (median change - 3 mmHg, 95% CI -6 to 0; p=0.06). There was no change in SV or CI.

In the 12 (52%) participants with PCWP greater than 15 mmHg before sotatercept, there were significant improvements in mPAP (median change -15 mmHg, 95% CI, -22 to -5, p<0.01), PCWP (median change -7 mmHg, 95% CI, -11 to -4, p<0.01), PVR (median change -2.7, 95% CI, -7.5 to -0.30; p=0.03), and PAC (median change 0.63, 95% CI, 0.28-0.97, p<0.01). There was no change in RAP, SV, or CI.

### Functional Class

Pre- and post-sotatercept FC are listed in Table S1. Functional class improved in 28 (58%) participants and did not worsen in any participants.

### Individual Participant Cases

Two cases had particularly noteworthy findings. One participant (“Participant A”) was a woman in her 50’s with a distant history of asthma who developed worsening dyspnea on exertion and cough over the past two years (FC III). Oxygen saturation at rest was 95% but would drop to 85% with exertion. PFTs demonstrated mildly reduced FVC, FEV1, and TLC, with marked reductions in DLCO (Table S2). Bronchodilators and inhaled corticosteroids provided no improvement in symptoms and oral prednisone only slightly improved her cough. CT chest revealed mild diffuse bronchial wall thickening and areas of gas trapping. RHC demonstrated precapillary PH and she was started on tadalafil without improvement in exercise capacity. Given the concern for potential ILD or pulmonary veno-occlusive disease, she underwent a surgical biopsy, which revealed bronchiolectasis with mucous plugging and intra-alveolar foamy macrophage accumulation (Figure 2A-D). Sotatercept was initiated for PAH. Within weeks, her exercise capacity improved (FC 2) and she had complete resolution of her cough. Repeat PFTs six weeks later demonstrated significant improvement in spirometry, lung volumes, and diffusing capacity (Table S2).

**Figure 2:**
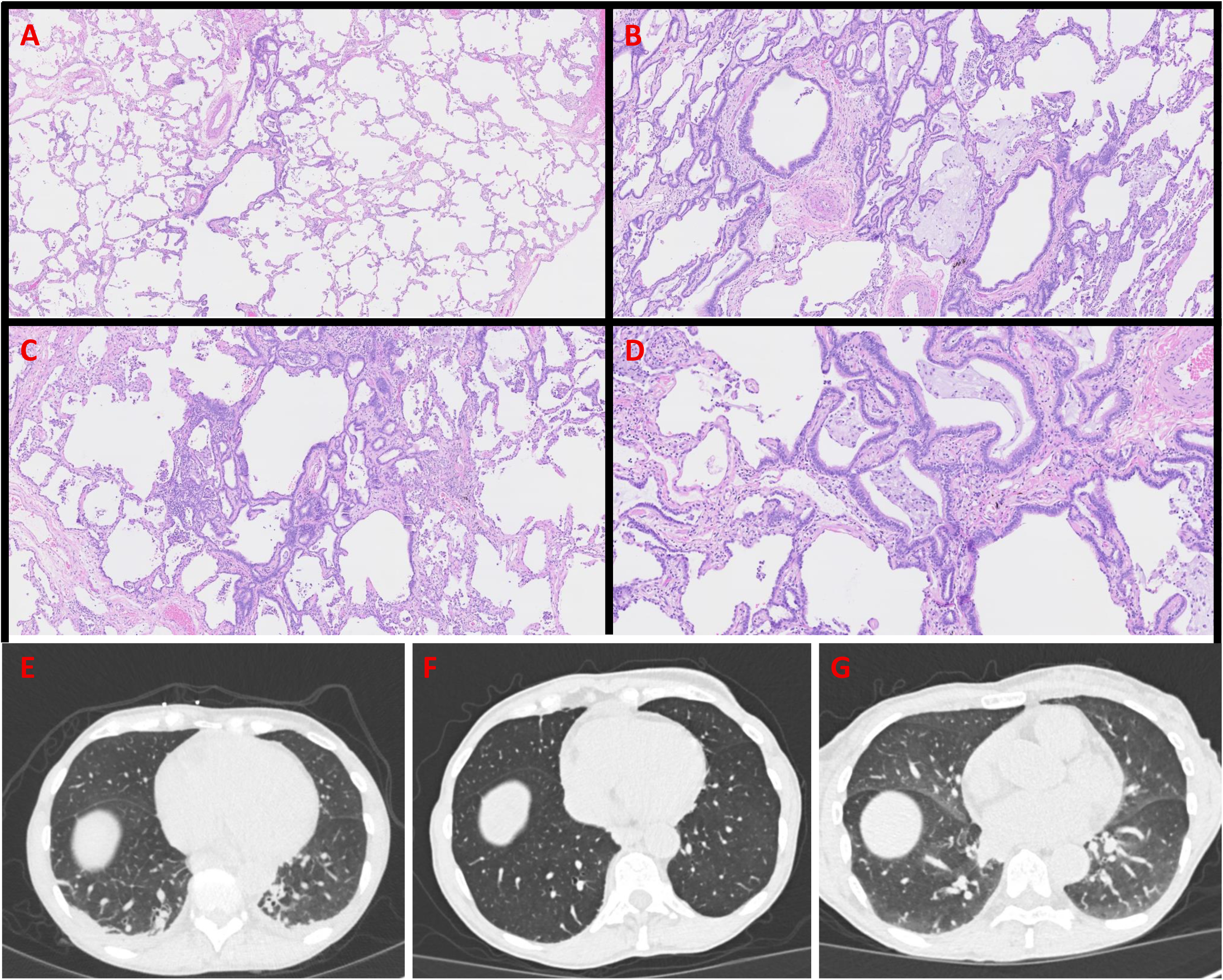
**(A-D)** Surgical lung biopsy specimen of a participant pre-sotatercept. **(A)** Peribronchiolar metaplasia and mild associated fibrosis. (H&E 20x) **(B)** Extensive peribronchiolar metaplasia with associated mild chronic inflammation, fibrosis and intra-luminal mucin accumulation. (H&E 50X) **(C)** Extensive peribronchiolar inflammation with associated fibrosis and chronic inflammation. Intra-luminal foamy macrophage accumulation is also present. (H&E 50x) **(D)** Peribronchiolar metaplasia with associated intra-luminal mucin and foamy macrophage accumulation. (H&E 100x) **(E-G)** Noncontrast chest CTs of a participant at the level of the right hemidiaphragm. **(E)** Pre-sotatercept inspiratory chest CT demonstrating bilateral dependent atelectasis. (**F&G)** Post-sotatercept inspiratory (**F**) and expiratory (**G**) chest CT scans seven weeks after sotatercept initiation, demonstrating resolution of bilateral dependent atelectasis.

Another participant (“Participant B”) was a woman in her 70’s with a history of idiopathic PAH diagnosed 16 years prior. She had been stable (FC II) on triple oral therapy (treprostinil, tadalafil, macitentan) for years but developed worsening dyspnea on exertion and hypoxemia over the past few months (FC III). Oxygen saturation was 86% while breathing room air. CT chest was unremarkable except for bilateral dependent atelectasis (Figure 2E). PFTs demonstrated normal FEV1 and FVC, but reduced FEV1/FVC and FEF25-75% (Table S3). Notably, TLC, FRC, and RV were elevated. She was initiated on sotatercept. Over the next few weeks, her exercise capacity improved (FC II) and oxygen saturation was 98% breathing room air. Repeat PFTs seven weeks later demonstrated significant improvement in spirometry, lung volumes, and diffusing capacity (Table S3). Repeat CT scan on the same day demonstrated resolution of bilateral dependent atelectasis (Figure 2F & 2G).

## DISCUSSION

In this retrospective cohort study of participants who received sotatercept for the treatment of severe precapillary PH, we note four major findings. First, while sotatercept was initiated with the intent of treating pulmonary vascular disease, there were unexpected improvements in lung function, particularly in markers of small airways function and hyperinflation. Second, a high percentage of participants (74%) who had body plethysmography had an RV/TLC greater than 100% predicted at baseline, despite the cohort including only one participant with COPD. Third, two major patterns of changes in lung volumes were noted after treatment with sotatercept, one consistent with a reduction in trapped air, and the other consistent with resolution of atelectasis. Fourth, in this cohort of precapillary PH associated with a variety of causes, sotatercept was associated with marked improvements in hemodynamics and FC.

Sotatercept was associated with increases in FEV1, FVC, FEF-25-75%, DLCO, and ERV, decreases in RV and RV/TLC, and no change in PEF, FRC, and TLC. This constellation of findings is consistent with improvement in small airways function and hyperinflation. The small airways are defined as airways less than two mm in diameter [23]. Small airways have a large total cross-sectional area at a given airway generation and contribute very little to airways resistance in health [24]. Because significant SAD is often necessary for FEV1 and/or FVC to be affected, FEF25-75% has been considered by some to be a more sensitive, albeit variable, index of small airways function. With FEF25-75%, flow rates are assessed at lower lung volumes where small airway diameter is smaller and elastic recoil pressure is less [25]. Median FEF25-75% in our study increased by 16% after sotatercept initiation. Of the 19 participants that had a reduction in FEF25-75%,15 (79%) had a marked increase in FVC (median increase 170 mL), indicating that small airway function likely improved in those participants as well. FEF25-75% may paradoxically fall in the setting of decreased small airways resistance when FVC increases because the flow rates are likely being measured at different absolute lung volumes [26]. There was no overall change in FEV1/FVC in our study. While 23 (48%) participants had a decline in FEV1/FVC, 20 (87%) had significant increases in FVC (median increase 225 mL), indicating improved airway function as the FEV1/FVC reduction was driven by an increase in FVC. PEF, a measurement that occurs near TLC and serves as a marker of large airway resistance, was unchanged in our study. DLCO improvements with sotatercept have been noted previously in patients with PAH; however, it is unclear if this improvement in our study or past studies is secondary to improvement in alveolo-capillary membrane conductance or pulmonary capillary blood volume [27].

The changes in lung volumes noted in the 22 participants that had body plethysmography both pre- and post-sotatercept appeared to follow two major patterns. When small airways are narrowed, three potential changes to lung volumes can theoretically occur (Figure 3): 1) Small airway occlusion can produce trapped air, which leads to an increase in RV, with associated increases in FRC and TLC (“trapped air” phenotype); 2) Small airway occlusion can result in atelectasis, which primarily leads to a reduction in ERV, with consequent decreases in FRC, TLC, and even RV (“atelectatic” phenotype); and 3) Small airway narrowing could lead to earlier small airway closure during each expiration, whereby decreases in ERV are converted to increases in RV. Given that after the initiation of sotatercept, participants who had a reduction in TLC also had a reduction in RV and changes in TLC correlated with changes in RV, sotatercept appears to improve lung function in some of the participants by reducing trapped air. Additionally, after the initiation of sotatercept, participants who had an increase in FRC also had an increase in ERV and changes in FRC correlated with changes in ERV. We therefore speculate that sotatercept also improves lung function in some participants by resolving atelectasis. Given there was no correlation between changes in ERV and changes in RV, it does not appear that sotatercept affects early airway closure. In one participant who appeared to have the “atelectatic” phenotype, her lung biopsy demonstrated bronchiolectasis and mucus plugging and body plethysmography after the initiation of sotatercept demonstrated an increase in FRC and ERV. In another patient who had the “trapped air” phenotype, including an elevated TLC (113% predicted), sotatercept was associated with a decrease in RV and TLC, along with resolution of dependent atelectasis. It is also possible that lung volume changes are not discrete phenotypes but represent a continuum or even a dynamic single process. Notably, pathologic specimens from patients with COPD demonstrate occlusion of the small airways with inflammatory exudates containing mucus [14].

**Figure 3:**
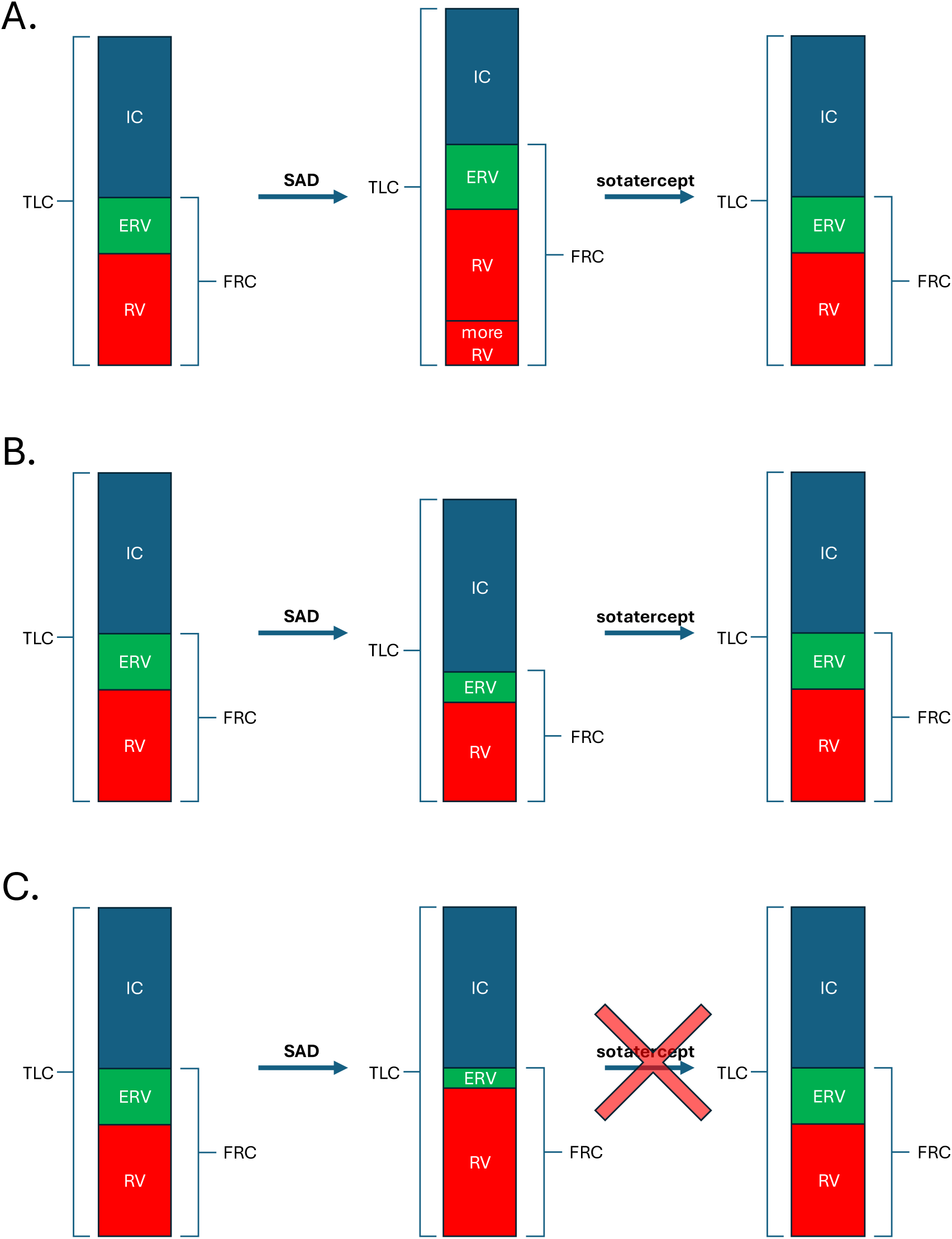
Theoretical models of changes in lung function with development of SAD and resolution with sotatercept treatment. **A.** “Trapped air” process. In the setting of small airway occlusion, additional air is trapped, leading to an increase in RV. Because this trapped air is also present at FRC and TLC, there are consequent increases in FRC and TLC. Sotatercept appears to improve this process. **B.** “Atelectatic” process. The airway segment becomes atelectatic in the setting of small airway occlusion, leading to a reduction in ERV, with consequent decreases in FRC and TLC. Because those lung segments also contain air that normally contribute to RV, there is a consequent decrease in RV as well. Sotatercept appears to improve this process **C.** “Early closure” process. Narrowing of small airways lead to earlier small airway closure, whereby decreases in ERV are converted to increases in RV. Sotatercept does not appear to improve this process. ERV: expiratory reserve volume, FRC: functional residual capacity, IC: inspiratory capacity, RV: residual volume, SAD: small airways disease, TLC: total lung capacity

The significant improvement in hemodynamics noted in our study could potentially be attributed to the improvements in SAD and hyperinflation. Increased intrathoracic pressure, whether it be from obesity or hyperinflation, can have significant effects on cardiopulmonary hemodynamics [28, 29]. With increases in intrathoracic pressure, both ventricles can be compressed, leading to increased pressures, smaller volumes, and impaired function [30–32]. The relationship between intrathoracic pressure and PCWP has been best characterized in obese patients, where post-capillary PH is frequently re-classified as precapillary PH (or even absence of PH) after accounting for intrathoracic pressure [33]. Patients with HFpEF frequently have abnormal PFTs [34], have high rates of SAD [10], and demonstrate limited ventilatory reserve during exercise [35]. Dynamic hyperinflation has been demonstrated in patients with HFpEF, the degree of which can explain at least some of the exercise-induced increase in PCWP [36]. Interestingly, improvement in SAD and hyperinflation may also explain why patients who received sotatercept in the STELLAR trial had a small but statistically significant decrease in left ventricular ejection fraction despite a decrease in PCWP [37]. Improvements in SAD lead to a reduction in hyperinflation, decreased intrathoracic pressure, and in turn, a reduction in PCWP and increase in LV afterload [38]. In our study, SAD and hyperinflation may have contributed to the elevated PCWP in some of the participants, and it is possible that the reduction in hyperinflation with sotatercept led to the decrease in PCWP. The results of the CADENCE trial (NCT04945460), which will assess the effect of sotatercept on CpcPH due to HFpEF, are greatly anticipated, though the study was not designed to assess for changes in lung function [21].

Randomized controlled trials of sotatercept for PAH have noted significant clinical improvements within as soon as several weeks [19, 20, 39]. Anecdotally, some participants in our study noted subjective improvement in exercise capacity in as soon as a few days after initiation sotatercept. Hyperinflation increases PVR by compression of alveolar capillaries [40–42]. In idiopathic PAH, inhaled salbutamol decreases PVR and increases CO, while improving markers of SAD [43]. It is possible that at least some of the reduction in PVR noted both in our study and in clinical trials for PAH are secondary to sotatercept’s potential effect on SAD. The speed of hemodynamic and clinical effect argues against vascular anti-remodeling as the sole, especially early, mechanism of sotatercept’s efficacy in PAH. We propose that sotatercept’s positive effect on SAD may have contributed to the rapid clinical improvements noted in the randomized controlled trials of sotatercept for PAH by improving both ventilatory efficiency and hemodynamics [19, 20, 37, 39].

Sotatercept’s purported major mechanism of action is activin-signaling inhibition in the pulmonary vasculature [22]. Activins drive macrophage activation, cytokine production (including IL-1β, IL-6, and TNF-α), and tissue infiltration [44]. In animal models of PAH, a sotatercept analogue reversed proinflammatory gene expression in macrophages and normalized pulmonary macrophage infiltration [45]. Activin signaling plays a significant role in the airways as well. Activin A levels are increased in the airway epithelium, airways smooth muscle cells, and alveolar macrophages of COPD patients, and cigarette smoke increases activin expression in the lungs [46]. In asthma, activin A promotes proliferation of airway smooth muscle and bronchial epithelial cells and contributes to hyperreactivity and airway remodeling [47, 48]. Serum activin A levels are higher in lung transplant patients that develop bronchiolitis obliterans syndrome [49]. It is therefore possible that sotatercept’s beneficial effect on SAD is also through activin-signaling inhibition.

The present study has several limitations. First, this study was a retrospective study with a small sample size that lacked a control group, and the timing of assessments (PFTs and RHCs) relative to sotatercept initiation was not standardized. This cohort was a heterogenous mix of “real world” patients with precapillary PH. While this heterogeneity may limit generalizability for each PH subgroup, the consistent increase in markers of SAD and hyperinflation in this heterogenous group is notable. As this was not a prospective, protocolized study, we cannot distinguish the effects of background pulmonary vasodilators on lung function and hemodynamics, nor primary respiratory treatments like bronchodilators. Many participants did not have body plethysmography or post-sotatercept hemodynamics, and thus, we could not assess the correlation between changes in lung volumes and changes in hemodynamics. While FC improved for many participants, formal exercise testing was not available. We did not directly measure intrathoracic pressure and did not include more direct measurements of SAD, like oscillometry, single or multiple breath nitrogen washout, functional CT imaging, or hyperpolarized magnetic resonance imaging.

In summary, in participants with precapillary PH from a variety of causes, sotatercept was associated with improvements in measures of SAD, hyperinflation, hemodynamics, and functional capacity. Future studies are needed to determine whether sotatercept could be a potential treatment for SAD.

## Data Availability

All data produced in the present study are available upon reasonable request to the authors

## Author’s contributions

H.D.P. concept, data gathering, data analysis, interpretation, drafting of the manuscript

J.Z. data gathering, data analysis, interpretation, revision of the manuscript

S.H-C. data gathering, interpretation, revision of the manuscript

S.S. data gathering, interpretation, revision of the manuscript

E.E. interpretation, revision of the manuscript

J.W. interpretation, revision of the manuscript

A.G.L. interpretation, revision of the manuscript

G.S. data gathering, interpretation, revision of the manuscript

H.S. interpretation, revision of the manuscript

L.R. interpretation, revision of the manuscript

B.S. interpretation, revision of the manuscript

M.B.B. data gathering, interpretation, revision of the manuscript

M.L. data analysis, interpretation, revision of the manuscript

C.A.P. interpretation, revision of the manuscript

C.E.V. interpretation, revision of the manuscript

M.P. interpretation, revision of the manuscript

---

**Figure S1:**
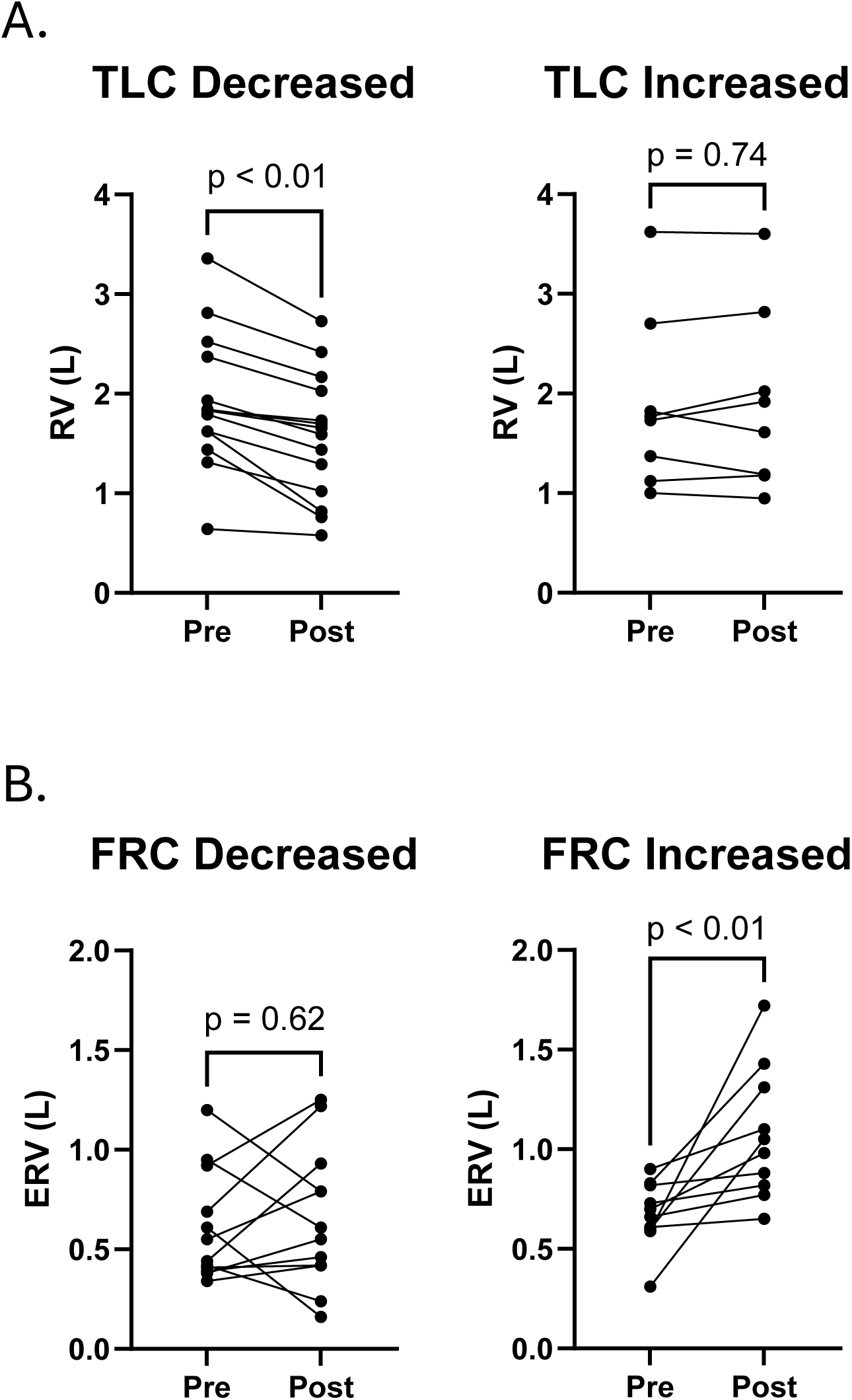
Lung volumes before and after sotatercept; specifically, RV and ERV changes relative to TLC and FRC changes. **A.** In participants who had a decrease in TLC, RV decreased (median RV change -340 mL, 95% CI, -490 to -220; p=0.001). In participants with an increase in TLC, there was no significant change in RV (median RV change +20 mL; 95% CI,-130 to 185; p=0.74). **B.** In participants who had an increase in FRC, all increases in ERV (median ERV increase 390 mL, 95% CI, 100-670, p<0.01). In participants who had a decrease in FRC, there was no change in ERV (median change in ERV 53 mL, 95% CI, -180 to 285; p=0.62). CI: confidence interval, ERV: expiratory reserve volume, FRC: functional residual capacity, RV: residual volume, TLC: total lung capacity

**Figure S2:**
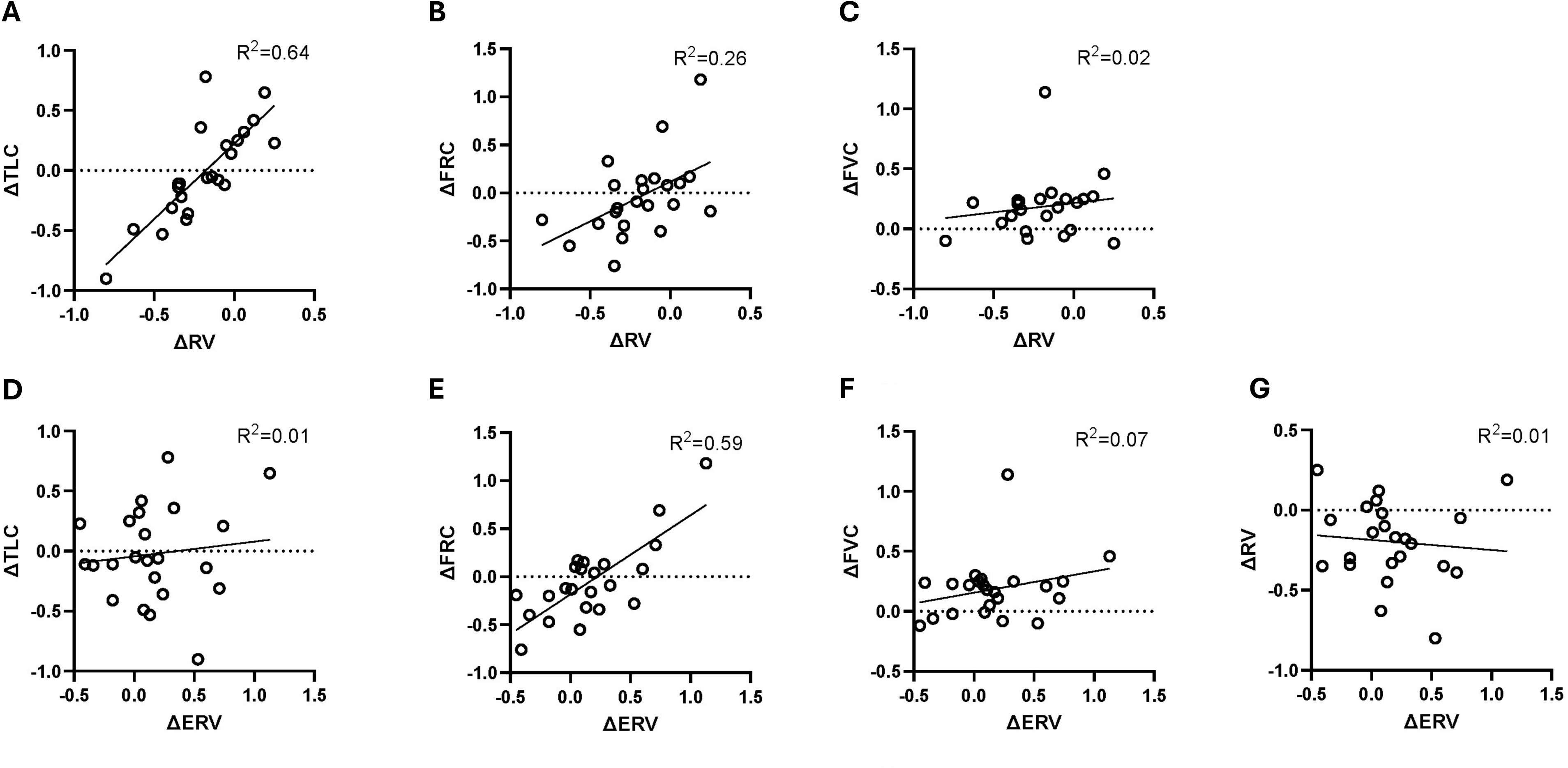
Correlations between changes in various lung volumes and capacities. There were strong correlations between changes in RV and changes in TLC, as well as changes in ERV and changes in FRC. There was a modest correlation between changes RV and changes in FRC. There was no correlation between changes in ERV and changes in RV. ERV: expiratory reserve volume, FRC: functional residual capacity, FVC: forced vital capacity, RV: residual volume, TLC: total lung capacity

**Figure S3:**
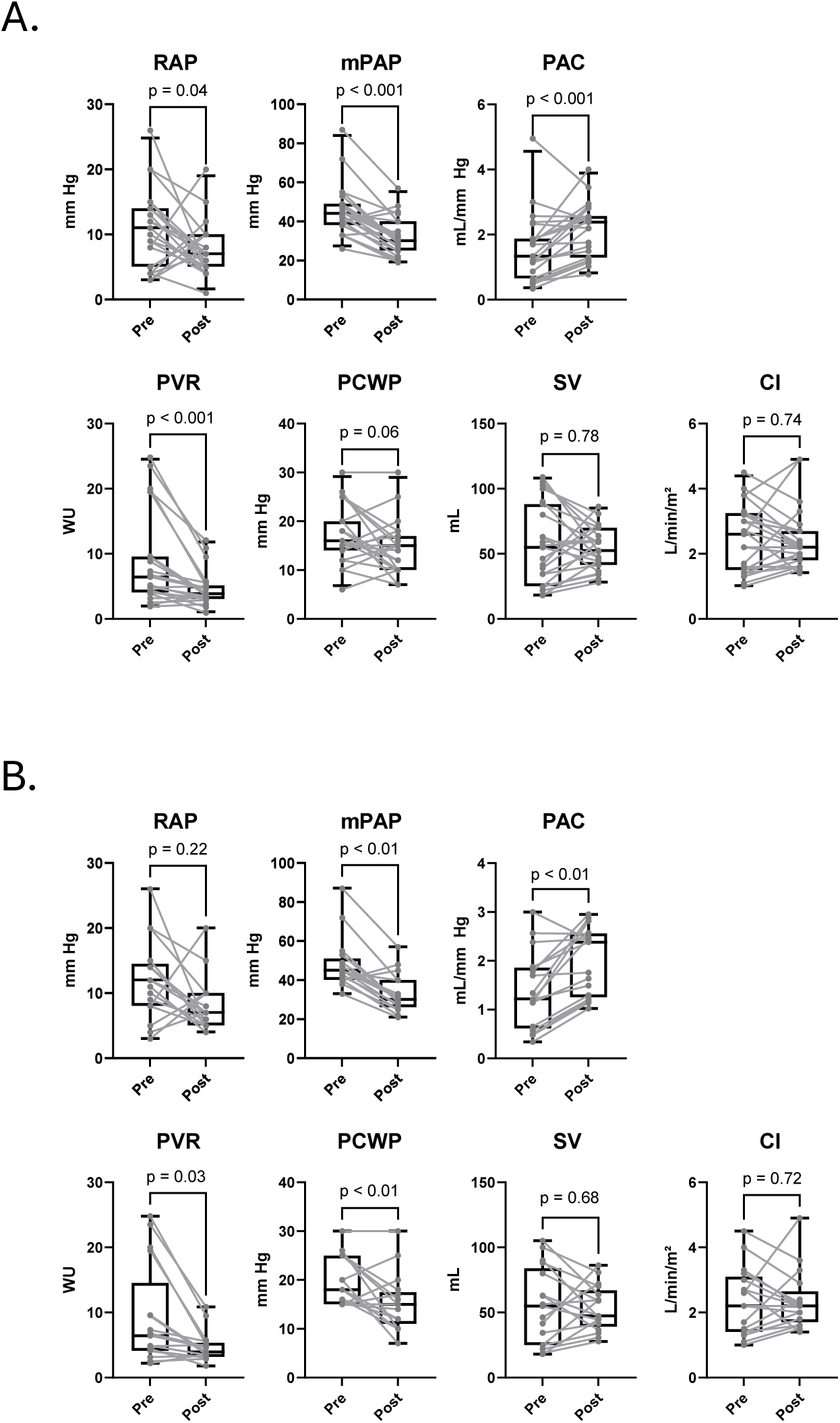
Changes in hemodynamics before and after administration of sotatercept. There were significant improvements in RAP, mPAP, PVR, PCWP, and PAC. **A.** Entire cohort. **B.** Participants with PCWP ≥ 15 mmHg. CI: cardiac index, mPAP: mean pulmonary artery pressure, PAC: pulmonary artery compliance, PCWP: pulmonary capillary wedge pressure, PVR: pulmonary vascular resistance, RAP: right atrial pressure, SV: stroke volume

**Table S1.**
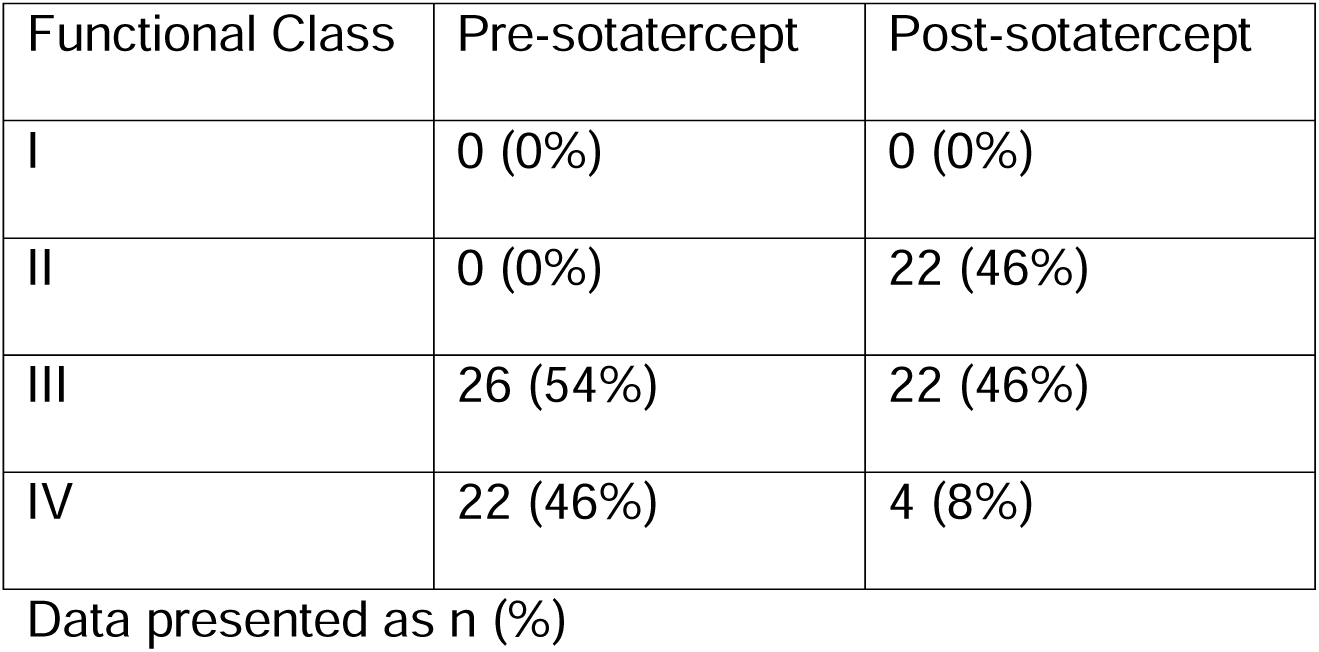
Changes in Functional Class.

| Functional Class | Pre-sotatercept | Post-sotatercept |
| --- | --- | --- |
| I | 0 (0%) | 0 (0%) |
| II | 0 (0%) | 22 (46%) |
| III | 26 (54%) | 22 (46%) |
| IV | 22 (46%) | 4 (8%) |

Data presented as n (%)

**Table S2.** “Participant A” Pulmonary Function Testing.

|  | Pre-Sotatercept | Post-Sotatercept | Change |
| --- | --- | --- | --- |
| FEV1, L | 1.46 (69%) | 1.67 (81%) | +0.21 |
| FVC, L | 1.82 (69%) | 2.07 (80%) | +0.25 |
| FEV1/FVC, % | 80% | 81% | +1% |
| FEF25-75%, L/s | 1.59 (79%) | 1.90 (96%) | +0.31 |
| DLCO, L/mmHg/min | 7.04 (39%) | 9.47 (53%) | +2.43 |
| TLC, L | 2.94 (64%) | 3.26 (72%) | +0.32 |
| FRC, L | 1.73 (72%) | 1.83 (78%) | +0.10 |
| RV, L | 1.12 (72%) | 1.18 (109%) | +0.06 |
| ERV, L | 0.61 (83%) | 0.65 (90%) | +0.04 |
| RV/TLC, % | 38% (115%) | 36% (90%) | -2% |

Data presented as measured value (percent predicted).
DLCO: diffusing capacity, ERV: expiratory reserve volume, FEF25-75%: forced expiratory flow between 25% and 75% of the vital capacity, FEV1: forced expiratory volume in one second, FVC: forced vital capacity, FRC: functional residual capacity, RV: residual volume, TLC: total lung capacity

**Table S3.** “Participant B” Pulmonary Function Testing.

|  | Pre-Sotatercept | Post-Sotatercept | Change |
| --- | --- | --- | --- |
| FEV1, L | 2.06 (93%) | 2.42 (110%) | +0.36 |
| FVC, L | 3.42 (119%) | 3.66 (127%) | +0.24 |
| FEV1/FVC, % | 60% | 66% | +6% |
| FEF25-75%, L/s | 0.90 (53%) | 1.30 (77%) | +0.40 |
| DLCO, L/mmHg/min | 12.9 (66%) | 13.3 (68%) | +0.4 |
| TLC, L | 5.94 (113%) | 5.83 (111%) | -0.11 |
| FRC, L | 3.72 (122%) | 2.96 (97%) | -0.86 |
| RV, L | 2.52 (114%) | 2.17 (98%) | -0.35 |
| ERV, L | 1.20 (162%) | 0.79 (107%) | -0.41 |
| RV/TLC, % | 42% (101%) | 37%(88%) | -5% |

Data presented as measured value (percent predicted).
DLCO: diffusing capacity, ERV: expiratory reserve volume, FEF25-75%: forced expiratory flow between 25% and 75% of the vital capacity, FEV1: forced expiratory volume in one second, FVC: forced vital capacity, FRC: functional residual capacity, RV: residual volume, TLC: total lung capacity

## Notes

**Sources of support:** C.E.V. R01HL174007; U54GM115677-09S3 (PD: Rounds); AHA 24IPA1275127

### Competing Interest Statement

H.D.P. advisory board for Merck
J.Z. none
S.H-C. none
S.S. none
E.E. none
J.W. none
A.G.L. none
G.S. none
H.S. none
L.R. none
B.S. none
M.B.B. none
M.L. none
C.A.P. none
C.E.V. consulting fees from Merck and Regeneron Pharmaceuticals; advisory boards for Janssen Pharmaceuticals and Merck; institution receives clinical trial support from Pulmovant, Gossamer Bio, Pfizer, Tenax Therapeutics, and Merck
M.P. consulting fees from Boehringer Ingelheim and United Therapeutics

### Funding Statement

C.E.V. received funding from: R01HL174007; U54GM115677-09S3 (PD: Rounds); AHA 24IPA1275127

### Author Declarations

The Mount Sinai Institutional Review Board gave ethical approval for this work (IRB approval 19-00263).

## References

1. Hassoun, P.M., Pulmonary Arterial Hypertension. N Engl J Med, 2021. 385(25): p. 2361–2376.

2. Trinkmann, F., et al., Small Airway Disease in Pulmonary Hypertension-Additional Diagnostic Value of Multiple Breath Washout and Impulse Oscillometry. J Clin Med, 2018. 7(12).

3. Fernandez-Bonetti, P., et al., Peripheral airways obstruction in idiopathic pulmonary artery hypertension (primary). Chest, 1983. 83(5): p. 732–8.

4. Meyer, F.J., et al., Peripheral airway obstruction in primary pulmonary hypertension. Thorax, 2002. 57(6): p. 473–6.

5. Rahaghi, F.N., et al., Evolution of Obstructive Lung Function in Advanced Pulmonary Arterial Hypertension. Am J Respir Crit Care Med, 2021. 204(12): p. 1478–1481.

6. Laveneziana, P., et al., Inspiratory muscle function, dynamic hyperinflation and exertional dyspnoea in pulmonary arterial hypertension. Eur Respir J, 2015. 45(5): p. 1495–8.

7. Trip, P., et al., Severely reduced diffusion capacity in idiopathic pulmonary arterial hypertension: patient characteristics and treatment responses. Eur Respir J, 2013. 42(6): p. 1575–85.

8. Olsson, K.M., et al., More on idiopathic pulmonary arterial hypertension with a low diffusing capacity. Eur Respir J, 2017. 50(2).

9. Kovacs, G., et al., Definition, classification and diagnosis of pulmonary hypertension. Eur Respir J, 2024. 64(4).

10. Nagumo, D., et al., Associations of Small-Airway Disease with Exercise Intolerance and Long-Term Outcomes in Patients with Heart Failure and Reduced or Preserved Ejection Fraction. Cardiology, 2024: p. 1–10.

11. Berigei, S.R., et al., Microscopic Small Airway Abnormalities Identified in Early Idiopathic Pulmonary Fibrosis In Vivo Using Endobronchial Optical Coherence Tomography. Am J Respir Crit Care Med, 2024. 210(4): p. 473–483.

12. Verleden, S.E., et al., Distinct Airway Involvement in Subtypes of End-Stage Fibrotic Pulmonary Sarcoidosis. Chest, 2021. 160(2): p. 562–571.

13. Quintero Santofimio, V., et al., Small Airways Obstruction and Mortality: Findings From the UK Biobank. Chest, 2024. 166(4): p. 712–720.

14. Hogg, J.C., et al., The nature of small-airway obstruction in chronic obstructive pulmonary disease. N Engl J Med, 2004. 350(26): p. 2645–53.

15. K Schwalbach, J.A., J Garry, H Nian, E Brittain, A Hemnes, Incidence and Clinical Associations with Phenotypic Drift from Pulmonary Arterial Hypertension to Combined Pre-and Post-capillary Pulmonary Hypertension. Chest Pulm, 2025, in press.

16. Hemnes, A.R., et al., Clinical Characteristics and Transplant-Free Survival Across the Spectrum of Pulmonary Vascular Disease. J Am Coll Cardiol, 2022. 80(7): p. 697–718.

17. Hoeper, M.M., et al., Idiopathic pulmonary arterial hypertension phenotypes determined by cluster analysis from the COMPERA registry. J Heart Lung Transplant, 2020. 39(12): p. 1435–1444.

18. Cassady, S.J. and M.T. Gladwin, The Mirage of Individualized Therapy in the United States: Barriers in Pulmonary Hypertension Due to Sickle Cell Disease & Sarcoidosis. Am J Respir Crit Care Med, 2025.

19. Humbert, M., et al., Sotatercept in Patients with Pulmonary Arterial Hypertension at High Risk for Death. N Engl J Med, 2025. 392(20): p. 1987–2000.

20. Hoeper, M.M., et al., Phase 3 Trial of Sotatercept for Treatment of Pulmonary Arterial Hypertension. N Engl J Med, 2023. 388(16): p. 1478–1490.

21. A Study of Sotatercept for the Treatment of Cpc-PH Due to HFpEF (MK-7962-007/ A011-16) (CADENCE). https://www.ClinicalTrials.gov identifier: NCT04945460. Updated August 8, 2025. Accessed September 22, 2025. https://www.clinicaltrials.gov/study/NCT04945460.

22. Yung, L.M., et al., ACTRIIA-Fc rebalances activin/GDF versus BMP signaling in pulmonary hypertension. Sci Transl Med, 2020. 12(543).

23. Ranga, V. and J. Kleinerman, Structure and function of small airways in health and disease. Arch Pathol Lab Med, 1978. 102(12): p. 609–17.

24. Macklem, P.T. and J. Mead, Resistance of central and peripheral airways measured by a retrograde catheter. J Appl Physiol, 1967. 22(3): p. 395–401.

25. Gelb, A.F. and N. Zamel, Simplified diagnosis of small-airway obstruction. N Engl J Med, 1973. 288(8): p. 395–8.

26. Cockcroft, D.W. and B.A. Berscheid, Volume adjustment of maximal midexpiratory flow. Importance of changes in total lung capacity. Chest, 1980. 78(4): p. 595–600.

27. Olsson, K.M., et al., Effects of sotatercept on lung diffusion capacity and blood gases in patients with pulmonary arterial hypertension. Eur Respir J, 2023. 62(2).

28. Behazin, N., et al., Respiratory restriction and elevated pleural and esophageal pressures in morbid obesity. J Appl Physiol (1985), 2010. 108(1): p. 212–8.

29. Haluszka, J., et al., Intrinsic PEEP and arterial PCO2 in stable patients with chronic obstructive pulmonary disease. Am Rev Respir Dis, 1990. 141(5 Pt 1): p. 1194–7.

30. Watz, H., et al., Decreasing cardiac chamber sizes and associated heart dysfunction in COPD: role of hyperinflation. Chest, 2010. 138(1): p. 32–8.

31. Barr, R.G., et al., Percent emphysema, airflow obstruction, and impaired left ventricular filling. N Engl J Med, 2010. 362(3): p. 217–27.

32. Kawut, S.M., et al., Cor pulmonale parvus in chronic obstructive pulmonary disease and emphysema: the MESA COPD study. J Am Coll Cardiol, 2014. 64(19): p. 2000–9.

33. Khirfan, G., et al., Impact of Esophageal Pressure Measurement on Pulmonary Hypertension Diagnosis in Patients With Obesity. Chest, 2022. 162(3): p. 684–692.

34. Huang, W.M., et al., The role of pulmonary function in patients with heart failure and preserved ejection fraction: Looking beyond chronic obstructive pulmonary disease. PLoS One, 2020. 15(7): p. e0235152.

35. G Cifci, D.B., B Borlaug, J Smith, Limited Ventilatory Reserve During Exercise in Heart Failure with Preserved Ejection Fraction. Chest Pulm, 2025. 3(3): p. 100138.

36. Leahy, M.G., et al., Heart-Lung Interactions in HFpEF: Dynamic Hyperinflation and Exercise PCWP. JACC Heart Fail, 2025. 13(8): p. 102523.

37. Souza, R., et al., Effects of sotatercept on haemodynamics and right heart function: analysis of the STELLAR trial. Eur Respir J, 2023. 62(3).

38. Buda, A.J., et al., Effect of intrathoracic pressure on left ventricular performance. N Engl J Med, 1979. 301(9): p. 453–9.

39. Nagaraj, S., et al., Hemodynamic effects of sotatercept administration in pulmonary hypertension-Insights from remote monitoring. J Heart Lung Transplant, 2025.

40. Hakim, T.S., R.P. Michel, and H.K. Chang, Effect of lung inflation on pulmonary vascular resistance by arterial and venous occlusion. J Appl Physiol Respir Environ Exerc Physiol, 1982. 53(5): p. 1110–5.

41. Schrijen, F.V., et al., Pulmonary vascular resistance rises with lung volume on exercise in obstructed airflow disease. Clin Physiol, 1989. 9(2): p. 143–50.

42. Harris, P., et al., The influence of the airways resistance and alveolar pressure on the pulmonary vascular resistance in chronic bronhcitis. Cardiovasc Res, 1968. 2(1): p. 84–92.

43. Spiekerkoetter, E., H. Fabel, and M.M. Hoeper, Effects of inhaled salbutamol in primary pulmonary hypertension. Eur Respir J, 2002. 20(3): p. 524–8.

44. Morianos, I., et al., Activin-A in the regulation of immunity in health and disease. J Autoimmun, 2019. 104: p. 102314.

45. Joshi, S.R., et al., Sotatercept analog suppresses inflammation to reverse experimental pulmonary arterial hypertension. Sci Rep, 2022. 12(1): p. 7803.

46. Verhamme, F.M., et al., Role of activin-A in cigarette smoke-induced inflammation and COPD. Eur Respir J, 2014. 43(4): p. 1028–41.

47. Kariyawasam, H.H., et al., Activin and transforming growth factor-beta signaling pathways are activated after allergen challenge in mild asthma. J Allergy Clin Immunol, 2009. 124(3): p. 454–62.

48. Gregory, L.G., et al., Overexpression of Smad2 drives house dust mite-mediated airway remodeling and airway hyperresponsiveness via activin and IL-25. Am J Respir Crit Care Med, 2010. 182(2): p. 143–54.

49. Veraar, C., et al., Potential novel biomarkers for chronic lung allograft dysfunction and azithromycin responsive allograft dysfunction. Sci Rep, 2021. 11(1): p. 6799.

